# Neuroprognostication via Spatially-Informed Machine Learning Following Hypoxic-Ischemic Injury

**DOI:** 10.1101/2025.01.08.25320190

**Authors:** John D. Lewis, Atiyeh A. Miran, Michelle Stoopler, Helen M. Branson, Ashley Danguecan, Krishna Raghu, Linh G. Ly, Mehmet N. Cizmeci, Brian T. Kalish

## Abstract

**Key Points:** *Question:* Can machine learning be used to reliably and accurately predict 18-month developmental outcomes from neonatal brain MRI following perinatal hypoxic-ischemic injury (HIE)?

*Findings:* In this cohort study we show that across cognitive, language, and motor domains, a machine learning model can predict 18-month developmental outcome scores for neonates with HIE with excellent accuracy, and can produce atlases of the brain regions responsible for developmental impairments.

*Meaning:* Machine learning can be used for automated neuroprognostication in HIE, and may not only produce accurate predictions, but also provide neuroanatomical information that may prove useful in the search for novel interventions.

**Importance:** Perinatal hypoxic-ischemic encephalopathy (HIE) is one of the most common causes of neonatal death and neurodevelopmental impairment worldwide. Accurate prognostication of developmental outcomes following perinatal HIE is an important component of family-centered and evidence-based care.

**Objective:** To utilize magnetic resonance imaging (MRI)-based radiomic measures together with machine learning to produce automated and objective predictions of developmental outcomes after perinatal HIE.

**Design:** This was a retrospective cohort study of infants born between January 2018 and January 2022 with HIE.

**Setting:** The data for this study were acquired at the neonatal neurocritical care unit of a quaternary care center based on the center’s institutional criteria for diagnosis and for the use of therapeutic hypothermia.

**Participants:** Neonates with a gestational age of ≥ 35 weeks and a diagnosis of neonatal encephalopathy.

**Exposure(s):** Therapeutic hypothermia, with a whole-body cooling system, was begun within 6 hours after birth and was continued for 72 hours.

**Main Outcome(s) and Measure(s):** Brain MRI data were acquired on postnatal day 4-5, after rewarming after completion of therapeutic hypothermia. At 18-months of age, developmental outcome measures were assessed with the Bayley Scales of Infant and Toddler Development. We extracted radiomic measures from the deep-gray matter structures and from 2224 cubic tiles across the entire brain, in multiple modalities, and provided these measures to an elastic-net penalized linear regression model to predict the 18-month developmental outcomes.

**Results:** MRI-based radiomic measures from 160 neonates were used in a 10-fold cross-validation framework to predict the 18-month Bayley outcome scores. Across cognitive, language, and motor domains, the mean correlation between the predicted outcomes and the observed outcomes was 0.947, and the mean coefficient of determination was 0.879.

**Conclusions and Relevance:** A machine learning model using MRI-based radiomic measures from infants with HIE can reliably predict their 18-month developmental outcomes with excellent accuracy across the full range of motor, cognitive, and language domains. In addition, our approach allowed us to map the predictor weightings into neuroanatomical space, producing atlases of the brain regions responsible for the developmental impairments; these may prove useful in the search for novel interventions.

## 1 Introduction

Hypoxic-ischemic encephalopathy (HIE) is caused by a disruption in blood flow or oxygenation during the perinatal period, and is a major cause of infant mortality and disability. Therapeutic hypothermia improves survival and neurodevelopmental outcome for neonates with HIE; but still, nearly half of these infants die or suffer neurodevelopmental impairments ^1–10^. Magnetic resonance imaging (MRI) is commonly used to assess the severity of brain injury in HIE and counsel families about the likelihood of neurodevelopmental sequelae ^3,11–18^. Injuries to deep gray-matter (DGM) structures, the posterior limbs of the internal capsule (PLIC), the cortex, and watershed zones have been associated with neurodevelopmental impairment after HIE ^11,12,15,17–19^. Current neuroprognostication relies on interpretation of MRI, which will vary with the quality of the imaging and the interpreter’s experience and visual acuity, and thus the accuracy of the clinical predictions of developmental outcome is limited. HIE may be associated with cerebral dysmaturation ^20^, but predicting this developmental sequelae is challenging.

To address this, we recently developed an approach for automated neuroprognostication using radiomic features in the DGM structures ^21^. Radiomics capture complex patterns that may fail to be seen with the naked eye ^22^, including features of the image intensity histogram; the relationships between image voxels; neighborhood gray-tone difference derived textures; and features of complex patterns. That work showed promise; however, it was not entirely satisfactory as it overlooked significant portions of the brain. In the present study, we expand the approach to use measures from the whole brain. We use the population-specific multi-contrast template from our earlier work ^21^, with its labels for the DGM structures and the PLIC; but additionally cover the entire brain with cubic tiles, each with a unique label. We then take those labels to each subject, and extract the measures under each label, in each modality of the data. Previously we used only T1-, T2-weighted data; here we additionally use diffusion-weighted data from which we obtain an apparent diffusion coefficient (ADC) map and a direction-averaged volume. This produced a very large number of measures — approximately 1.9 million, with the inclusion of the scanner model as an interaction term — so we use an elastic-net penalized linear regression model to relate those measures to the developmental outcomes ^23^. We supply the radiomic measures to this elastic-net penalized linear regression model and use it to predict the 18-month developmental outcomes. We hypothesized that this advanced machine-learning approach, using spatial radiomics of the entire brain, would yield high accuracy in predicting the developmental outcomes in infants with perinatal HIE. Moreover, our approach weights the contribution of each of the radiomic measures that contribute to the predictions, indicating the prognostic value of the brain regions from which they were extracted; thus we are able to produce atlases of the brain regions that contribute to each domain of developmental impairments.

## 2 Methods

### 2.1 Study Cohort, Clinical and Laboratory Parameters

This retrospective cohort study was conducted at the neonatal neurocritical care unit of the Hospital for Sick Children in Toronto, Canada. The study protocols (REB:1000064940, 1000079302) were reviewed and approved by the Institutional Research Ethics Board, and informed consent was waived. The subjects are neonates born between January 2018 and January 2022, with a gestational age of ≥ 35 weeks with HIE. Neonates underwent therapeutic hypothermia if they met institutional criteria for mild, moderate, or severe neonatal encephalopathy.

Therapeutic hypothermia, with a whole-body cooling system, was begun within 6 hours after birth and was continued for 72 hours, unless discontinued early due to clinical contraindications. An MRI was acquired at approximately 4 days after birth, after completion of therapeutic hypothermia. The acquisition protocol produced (i) a 3D T1-weighted volume, (ii) T2-weighted volumes in multiple orientations, and (iii) diffusion-weighted data. In instances where issues, *e.g.* motion, resulted in poor scan quality, the scan was repeated to obtain usable data. Basic demographic, clinical, and laboratory (biochemical and encephalopathy) measures were obtained from the hospital’s electronic medical records. The T1- and T2-weighted MRI-data from all infants, if acceptable, were used to construct a population-specific multi-contrast template. The demographic, clinical, and laboratory data for those infants are presented in ^21^, as well as the scan protocol for the T1- and T2-weighted data, the processing methods for those data, and the methods for construction of the template. Here, we used the same methods to process the T1- and T2-weighted data, and that same template; but in addition to the labels on the DGM structures and the PLIC, we covered the brain with cubic tiles (7mm iso; 2224 tiles), each with a unique label. The population-specific template with these labels is shown in Supplementary eFigure 1. The diffusion-weighted data were processed into an apparent diffusion coefficient (ADC) map and a direction-averaged maximum b-value volume, each of which was considered a modality.

For those infants for whom their neonate T1-, T2-, and diffusion-weighted data were acceptable, and for whom we also had their 18-month behavioral outcome data, this template was then overlaid on their MRI-data, and radiomic measures were extracted from each modality for each label. There are 107 radiomic measures associated with each modality for each label; thus, with the scanner model as an interaction term, there are approximately 1.9 million MRI-based potential predictors. Our use of the elastic-net penalized linear regression machine learning model allowed this huge set of potential predictors to be reduced to the best set of predictors for each developmental outcome. Elastic-net penalized linear regression balances the approach to the regularization of the coefficients used in Ridge regression, that keeps the coefficients small, but keeps all variables in the model, with the approach used in Lasso regression, that allows some coefficients to go to zero ^23^. The elastic-net model aims for a balance which allows for learning a sparse model where few of the weights are non-zero, and the coefficients are generally kept from becoming large. We used a nested 10-fold cross-validation framework to ensure that our results generalize. For each outer fold, the elastic-net model was fitted on the training data, and predictions made for the testing data; an inner 10-fold cross-validation loop determined the hyper-parameters that yielded the best balance between the two approaches to regularization. The performance of the elastic-net penalized linear regression model was assessed via three evaluation metrics: the correlation coefficient (R) between the predicted and observed outcomes; the coefficient of determination (R^2^) — an estimate of the proportion of variance in the observed outcomes that can be explained by the predictors; and the mean absolute error (MAE) of the predictions.

We assessed the predictions based on only these MRI-based measures, since our goal was to show that the MRI-based measures alone yield good predictions for each of the Bayley-III outcome measures. Basic demographic and laboratory (including biochemical and clinical encephalopathy) measures were obtained from the electronic medical records. The prediction analyses utilize the subset of the infants for whom we had both good quality MRI and 18-month outcome assessments. Many of the infants had not yet completed their 18-month assessment; a number of infants could not complete their 18-month assessment due to COVID-19; a few families moved away before their infant turned 18 months old; and a small number of infants did not survive. One infant with severe CP and global developmental delay could not be assessed; a percentile score of 1 was assigned across all Bayley outcome domains for this infant.

## 3 Results

Supplementary eTables 1 through 5 provide the details of the demographic and laboratory data in the context of each of the prediction analyses.

### 3.1 Cognitive outcomes

The demographic and laboratory data for the infants for whom we had both all MRI modalities and Bayley-III cognitive outcome scores (n=168) are presented in Supplementary eTable 1. Of these infants, 62.5% had normal brain MRIs, 19.6% had predominantly white-matter/watershed injuries, 9.5% had predominantly DGM injuries, and 8.3% had near-total injury. For these infants, our predictions were strongly correlated with the cognitive outcomes (r:0.946, 95% CI[0.927, 0.960]) and explained much of the variance in the observed outcomes (r^2^:0.894). These predictions are plotted in Figure 1. Most of the predictors (89%) came from outside of the DGM structures; but also most of the DGM structures were predictors. The top predictors were in the left globus pallidus, the left superior temporal lobe, the right cerebellum, and the left and right superior frontal lobes. The maps of these predictors are presented in Figure 1. These maps show the sum of the coefficients for the predictors from all modalities, both summed across modalities, and for each modality separately.

**Figure 1:**
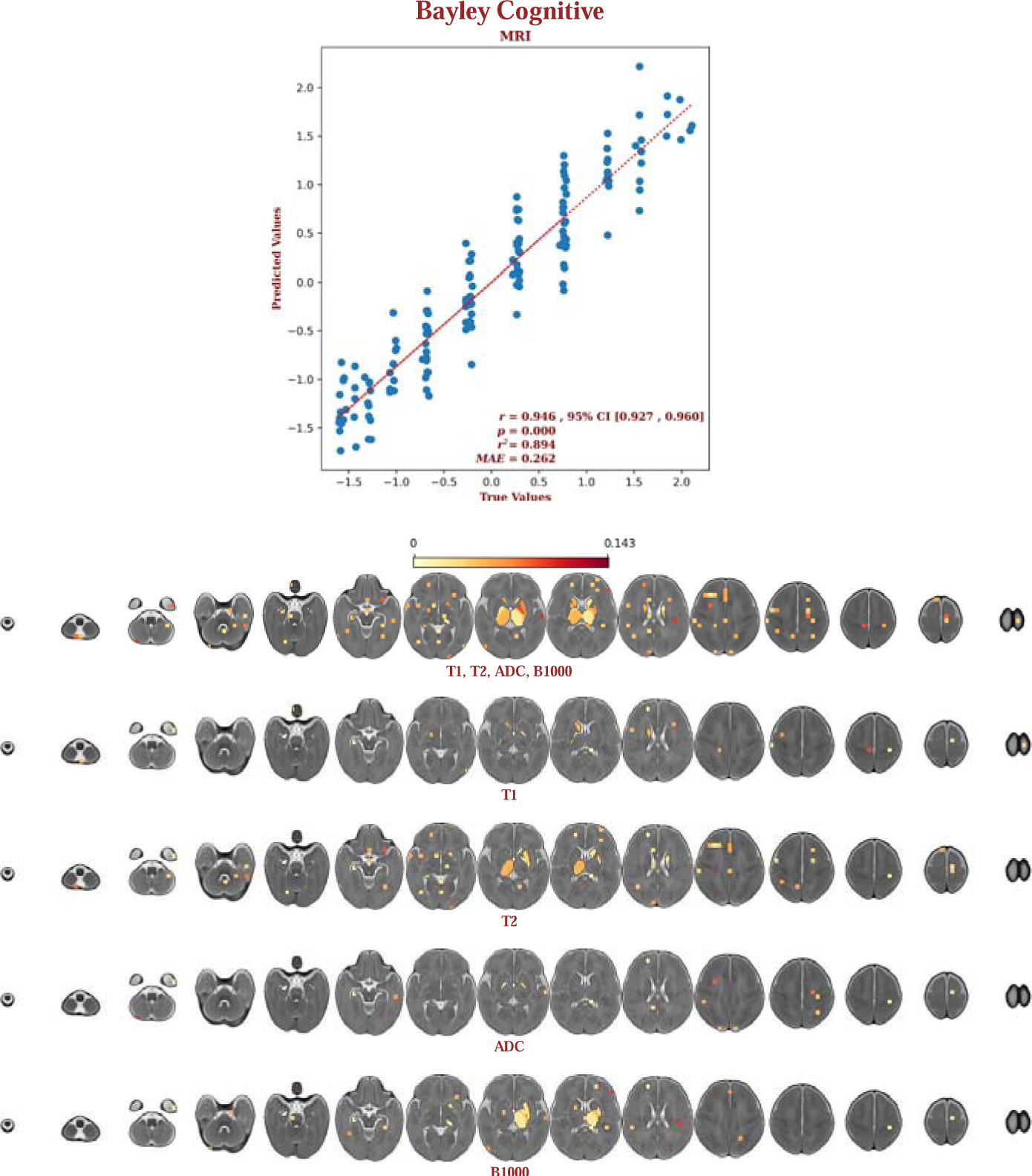
The elastic-net regression results for the Bayley cognitive scores. The predictors are shown below on axial slices in radiological view; the sum of the coefficients for each label is shown on the yellow-red scale. The top row of predictors shows the sum of the coefficients for the predictors from all modalities. The remaining rows show the sum of the coefficients for the predictors from each of the T1, the T2, the ADC map, and the direction-averaged maximum b-value volume. Note that most of the deep gray-matter structures are predictors, with the left pallidus being particularly prominent. But note also that predictors are distributed throughout the brain.

### 3.2 Expressive language outcomes

The demographic and laboratory data for the infants for whom we had both all MRI modalities and Bayley-III expressive language outcome scores (n=153) are presented in Supplementary eTable 2. Of these infants, 64.9% had normal brain MRIs, 17.6% had predominantly white-matter/watershed injuries, 9.5% had predominantly DGM injuries, and 8.1% had near-total injury. For these infants, our predictions were strongly correlated with the expressive language outcomes (r:0.931, 95% CI [0.905, 0.950]) and explained much of the variance in the observed outcomes (r^2^:0.851). These predictions are plotted in Figure 2. Most of the predictors came from outside of the DGM structures; 25% came from the DGM structures or PLIC, with almost all of those coming from the right hemisphere. The top predictors were in the right caudate, the right hippocampus, amygdala, and PLIC, and in cubic tiles in the anterior left superior frontal lobe and the superior temporal lobe. The maps of these predictors are presented in Figure 2. These maps show the sum of the coefficients for the predictors from all modalities, both summed across modalities, and for each modality separately.

**Figure 2:**
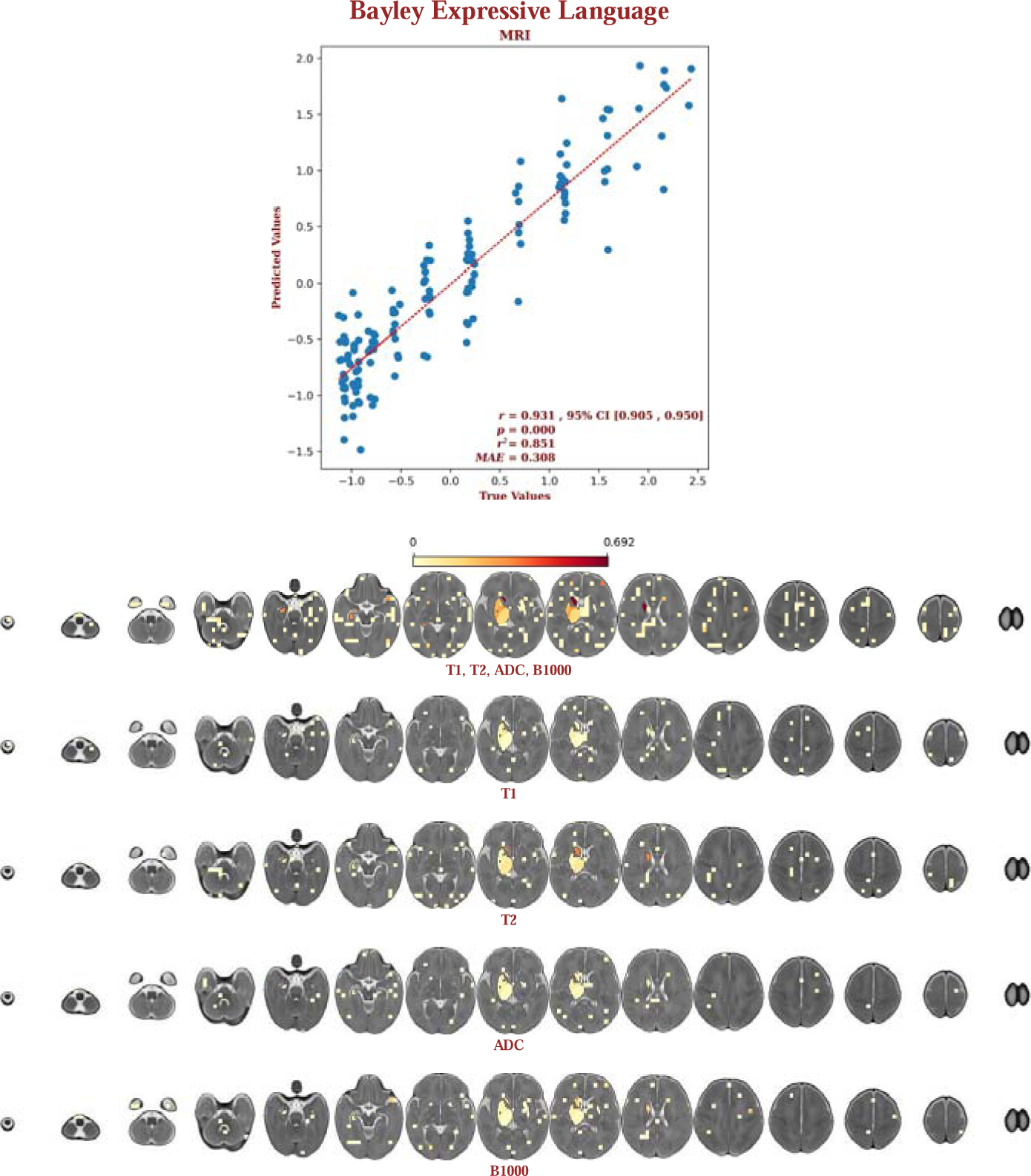
The elastic-net regression results for the Bayley expressive language scores. The predictors are shown below on axial slices in radiological view; the sum of the coefficients for each label is shown on the yellow-red scale. The top row of predictors shows the sum of the coefficients for the predictors from all modalities. The remaining rows show the sum of the coefficients for the predictors from each of the T1, the T2, the ADC map, and the direction-averaged maximum b-value volume. Note that the predictors are distributed throughout the brain, but are particularly prominent in the deep gray-matter structures of the right hemisphere.

### 3.3 Receptive language outcomes

The demographic and laboratory data for the infants for whom we had both all MRI modalities and Bayley-III receptive language outcome scores (n=153) are presented in Supplementary eTable 3. Of these infants, 64.9% had normal brain MRIs, 17.6% had predominantly white-matter/watershed injuries, 9.5% had predominantly DGM injuries, and 8.1% had near-total injury. For these infants, our predictions were strongly correlated with the receptive language outcomes (r:0.959, 95% CI [0.943, 0.970]) and explained much of the variance in the observed outcome data (r^2^:0.912). These predictions are plotted in Figure 3. Most of the predictors came from outside of the DGM structures; only 1% came from the DGM structures. The top predictors were in tiles near the left parieto-occipital junction, at the edge of the right thalamus, and spanning the left and right middle parietal lobes. The maps of these predictors are presented in Figure 3. These maps show the sum of the coefficients for the predictors from all modalities, both summed across modalities, and for each modality separately.

**Figure 3:**
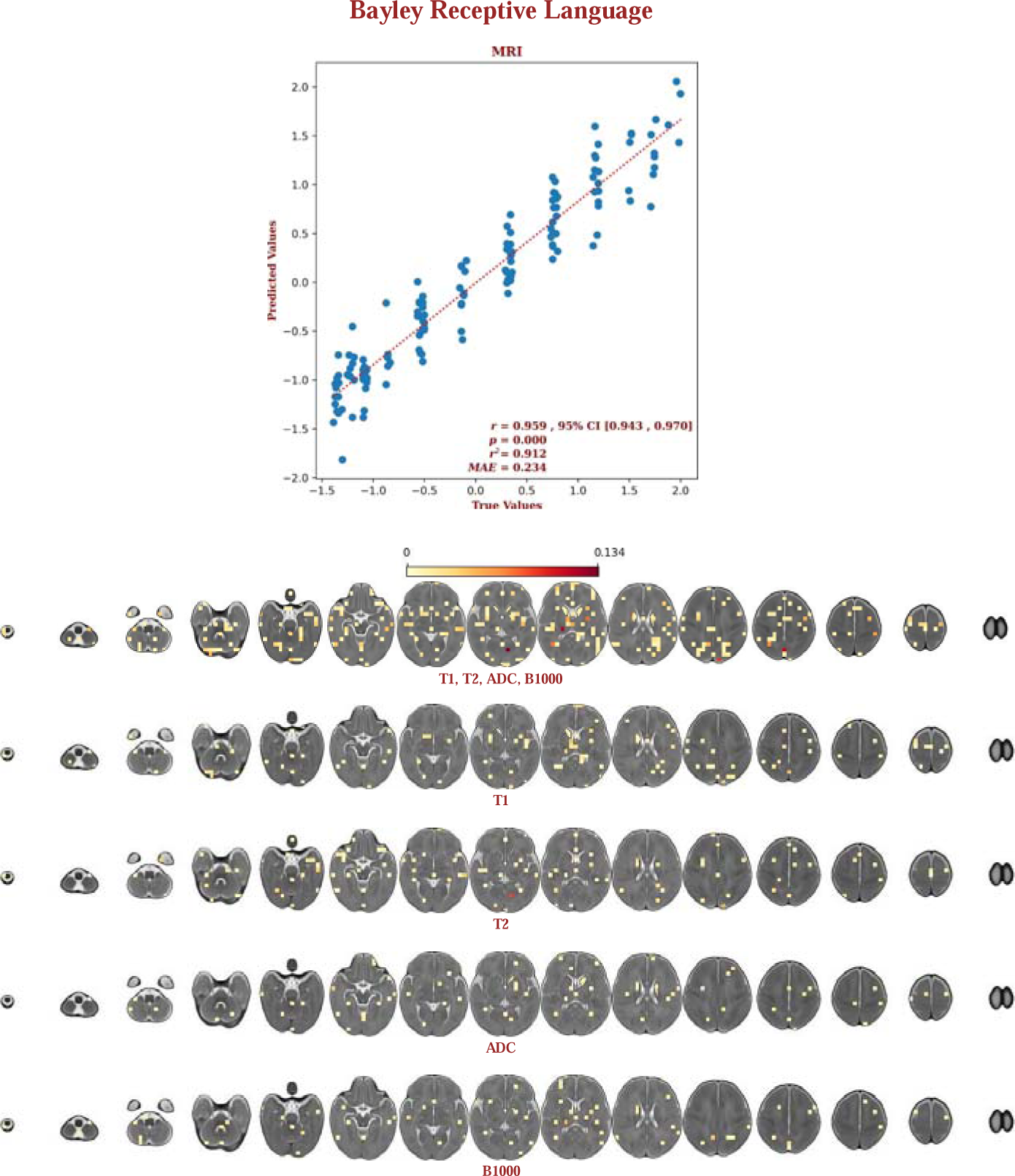
The elastic-net regression results for the Bayley receptive language scores. The predictors are shown below on axial slices in radiological view; the sum of the coefficients for each label is shown on the yellow-red scale. The top row of predictors shows the sum of the coefficients for the predictors from all modalities. The remaining rows show the sum of the coefficients for the predictors from each of the T1, the T2, the ADC map, and the direction-averaged maximum b-value volume. Note that the predictors are distributed throughout the brain, but with little involvement of the deep gray-matter structures.

### 3.4 Gross motor outcomes

The demographic and laboratory data for the infants for whom we had both all MRI modalities and Bayley-III gross motor outcome scores (n=161) are presented in Supplementary eTable 4. Of these infants, 63.2% had normal brain MRIs, 20.0% had predominantly white-matter/watershed injuries, 10.3% had predominantly DGM injuries, and 6.5% had near-total injury. For these infants, our predictions were strongly correlated with the gross motor outcomes (r:0.944, 95% CI [0.924, 0.959]) and explained much of the variance in the observed outcome data (r^2^:0.845). These predictions are plotted in Figure 4. Most of the predictors came from outside of the DGM structures; less than 1% came from the DGM structures or PLIC. As might be expected, there were predictors in the cerebellum and in motor regions of the cortex, but predictors were also widely distributed throughout the white-matter and the cortex. The top predictors were in the orbito-frontal cortex, and at the outer edge of the central sulcus near the junction of the superior and middle gyrus. The maps of these predictors are presented in Figure 4. These maps show the sum of the coefficients for the predictors from all modalities, both summed across modalities, and for each modality separately.

**Figure 4:**
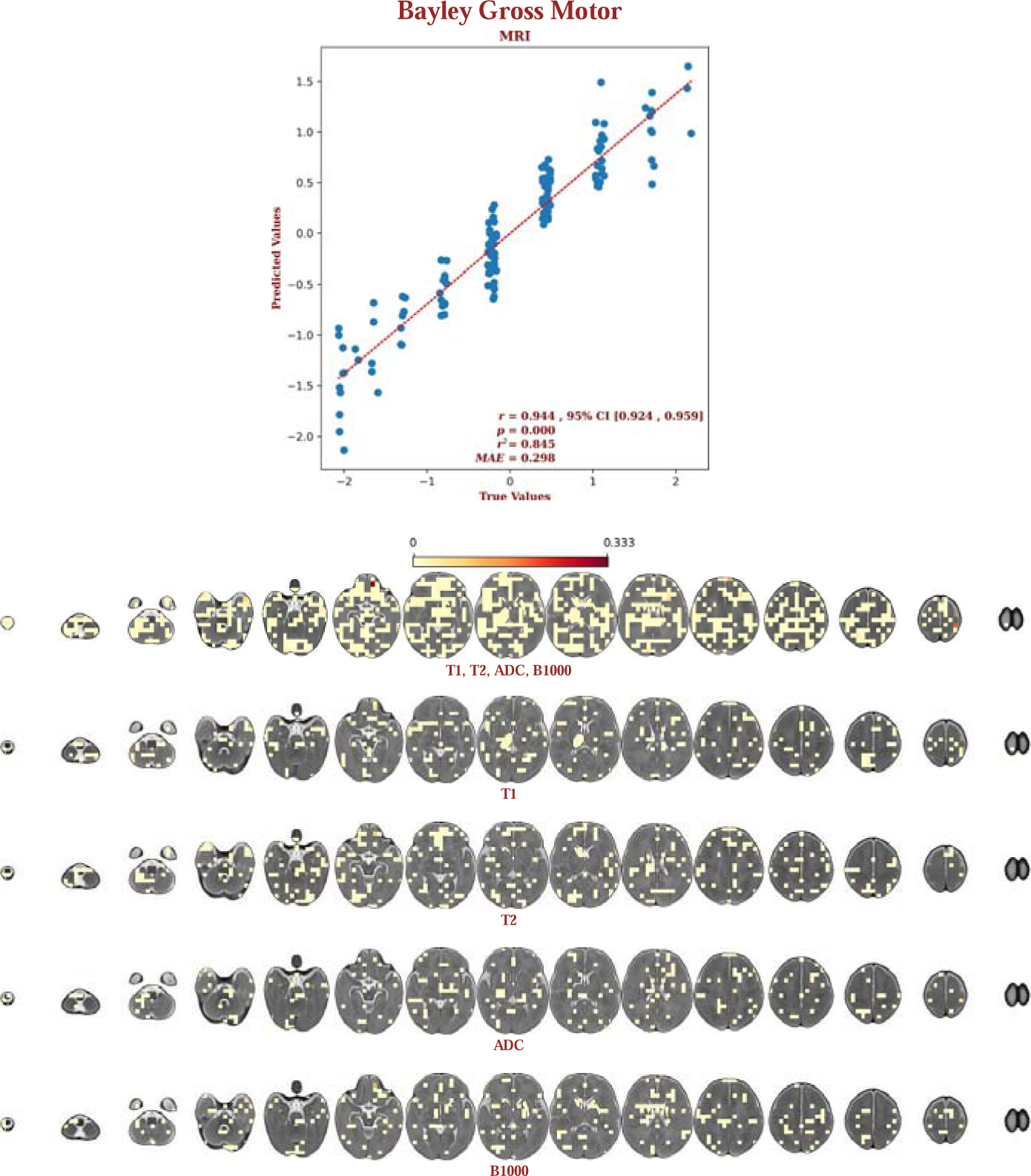
The elastic-net regression results for the Bayley gross motor scores. The predictors are shown below on axial slices in radiological view; the sum of the coefficients for each label is shown on the yellow-red scale. The top row of predictors shows the sum of the coefficients for the predictors from all modalities. The remaining rows show the sum of the coefficients for the predictors from each of the T1, the T2, the ADC map, and the direction-averaged maximum b-value volume. Note that the predictors are distributed throughout the white matter, the deep gray-matter structures, the cortex, and the cerebellum.

### 3.5 Fine motor outcomes

The demographic and laboratory data for the infants for whom we had both all MRI modalities and Bayley-III fine motor outcome scores (n=165) are presented in Supplementary eTable 5. Of these infants, 62.3% had normal brain MRIs, 20.1% had predominantly white-matter/watershed injuries, 10.1% had predominantly DGM injuries, and 7.5% had near-total injury. For these infants, our predictions were strongly correlated with the fine motor outcomes (r:0.955, 95% CI [0.938, 0.967]) and explained much of the variance in the observed outcome data (r^2^:0.893). These predictions are plotted in Figure 5. Most of the predictors came from outside of the DGM structures; less than 1% came from the DGM structures or PLIC. There were predictors in the cerebellum and in motor regions of the cortex, but predictors were also widely distributed throughout the white-matter and the cortex. The top predictors were in tiles that overlapped with the cerebellum, the brainstem, the corpus callosum, and the medial cortex at the parieto-occipital junction. The maps of these predictors are presented in Figure 5. These maps show the sum of the coefficients for the predictors from all modalities, both summed across modalities, and for each modality separately.

**Figure 5:**
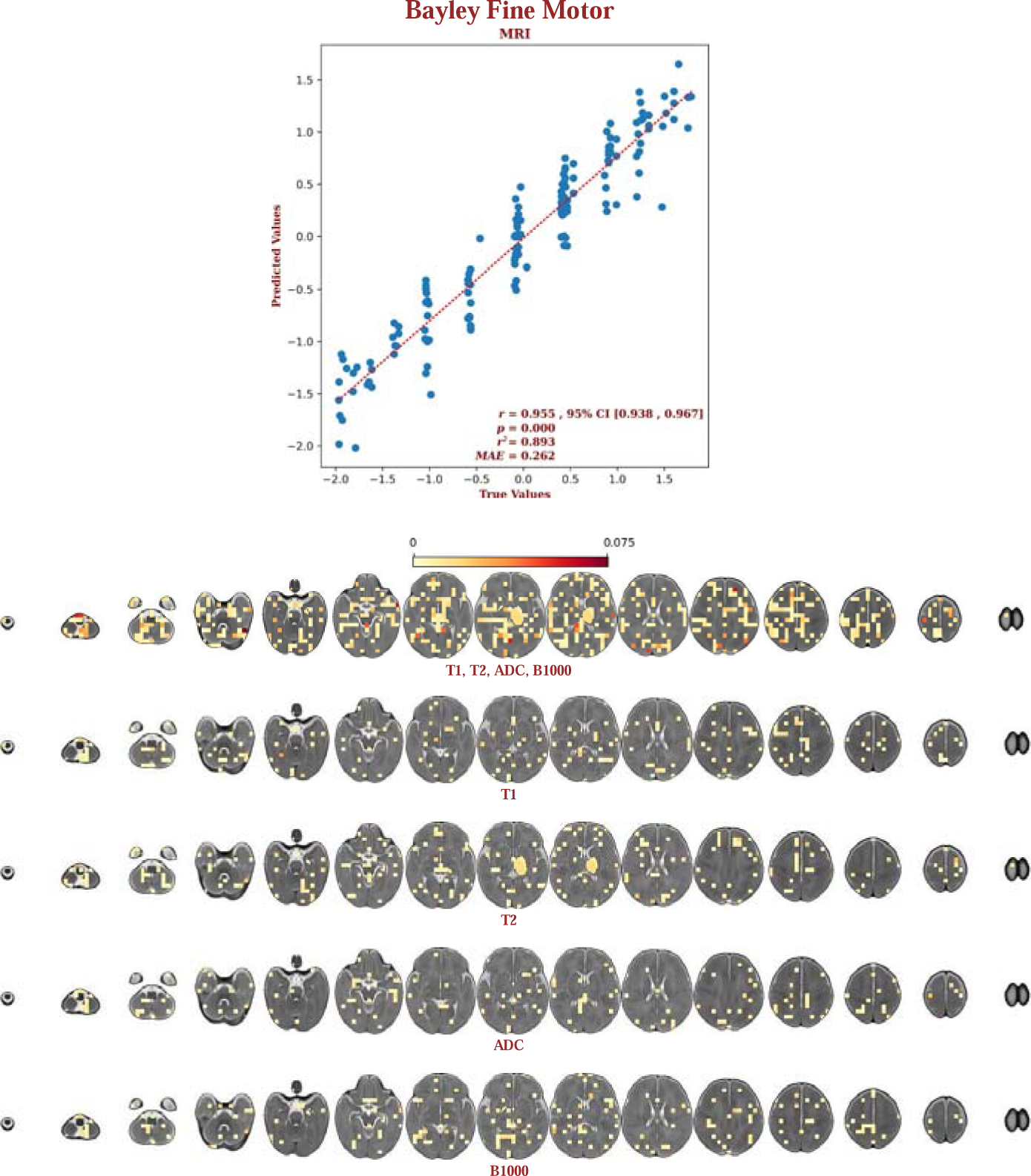
The elastic-net regression results for the Bayley fine motor scores. The predictors are shown below on axial slices in radiological view; the sum of the coefficients for each label is shown on the yellow-red scale. The top row of predictors shows the sum of the coefficients for the predictors from all modalities. The remaining rows show the sum of the coefficients for the predictors from each of the T1, the T2, the ADC map, and the direction-averaged maximum b-value volume. Note that the predictors are distributed throughout the white matter, the deep gray-matter structures, the cortex, and the cerebellum.

## 4 Discussion

Current approaches to neuroimaging-based prognostication rely on human expertise, and are thus subjective and have limited accuracy ^17,24–27^. We have described a new automated approach that is highly accurate, with a mean correlation of 0.947 between the predicted and observed outcomes across domains, and a mean coefficient of determination of 0.879. Importantly, the model explained variance in the observed outcomes to approximately the same degree regardless of severity, indicating that neonatal MRI has predictive power for the full spectrum of outcomes in HIE. Moreover, our approach identified the neuroimaging features that drove those predictions, allowing us to create atlases of the predictors of each of the developmental outcomes. This may provide valuable insight into the patterns of injury responsible for developmental impairments.

Importantly, the predictor atlases vary between each of the developmental outcome domains. The atlases for cognitive outcomes and for expressive language outcomes have predictors throughout the brain, but with prominent predictors in the DGM structures, perhaps in partial agreement with some previous research, e.g. ^11,28^, which associated damage to the basal-ganglia and thalamus with cognitive and language impairments. But notably, for cognitive outcomes, the predictors in the DGM structures are bilateral, whereas for the expressive language outcomes, the predictors in the DGM structures are concentrated in the right hemisphere. The concentration of predictors in the right-hemisphere DGM structures for the expressive language outcomes may reflect the innate asymmetry of brain structures involved in learning the prosodic structure of language versus its syntactic structure ^29–31^. Aspects of language more related to prosody are right lateralized ^29–32^. The lack of predictors in the DGM structures for the receptive language outcomes seems consistent with the fact that these results are from MRI data from neonates who have had essentially no exposure to language outside the womb.

The atlases for both the gross and fine motor outcomes have predictors, unsurprisingly, in the cerebellum, but somewhat surprisingly, in almost all other regions of the brain, as well. This lack of involvement of the DGM structures is in partial conflict with previous findings given that such damage has been associated with motor impairments, including cerebral palsy ^11^, though cerebral palsy has also been associated with white-matter damage ^33,34^. The widely distributed predictors throughout the brain in the atlases for each of the outcomes is in more general agreement with previous research ^28^, though the atlases provided here add specificity to those claims, and differentiate the relatively sparse distribution of predictors for cognitive outcomes from the more dense distribution of predictors for motor outcomes.

This study has several limitations that need to be taken into account when interpreting the results. First, the data were acquired at a single center, and were based on that center’s institutional criteria for diagnosis and for the use of therapeutic hypothermia. Notably however, our study population included a large number of neonates with mild encephalopathy, and whose brain MRIs appeared normal. We believe that inclusion of a broad spectrum of injury patterns demonstrates the ability of our approach to prognosticate across the full spectrum of severity. However, this may not generalize, and thus our findings require replication in samples acquired at centers with different clinical practices and patient populations. Our analyses may also have been impacted by the use of different MRI scanners with different field strengths, even though the scanner model was included as an interaction term. The analyses might also have been negatively impacted by the suboptimal slice thickness used for the T2- and diffusion-weighted data. However, it should be noted that despite this limitation, the radiomics features chosen as predictors by the elastic-net model were as likely to come from the T2- or diffusion-weighted data as from the T1-weighted data.

In conclusion, we have demonstrated that machine learning, using radiomics, has the potential to accurately predict 18-month outcomes in infants with perinatal HIE across the full spectrum of outcomes, in all domains, i.e. cognitive, language, and motor. Additionally, our methods have yielded atlases of the predictors for each of the outcome measures for each modality, which may provide insight into the nature of the brain injuries that drive the observed impairments, and guidance for future studies that strive to determine in more detail the nature of the injuries. Finally, we note that this study considered the MRI-data alone in order to investigate the prognostic value of that data; including the demographic, clinical, and laboratory data may yield still better results.

## Data availability

Our multi-contrast population-specific neonatal brain MRI template and the labels for the cubic tiling, the deep gray-matter structures, and the PLIC, can be found here: https://gin.g-node.org/johndlewis/HIE3/Template/; the scripts used to process the data can be found here: https://gin.g-node.org/johndlewis/HIE3/Tools; and the linear regression model can be found here: https://gin.g-node.org/johndlewis/HIE3/Models/.

## Acknowledgements

This research was enabled in part by support provided by the TD Bank Group Charity Classic Golf Tournament and in part by the Digital Research Alliance of Canada (alliancecan.ca). Drs. Mehmet N. Cizmeci and Linh G. Ly received support from Dr. Karen Pape Program in Neuroplasticity for neuroprognostication and neurodevelopmental research. Our thanks to the Neonatal Neurodevelopmental Follow-Up Clinic team for their efforts to capture our outcome data, and to Vladimir S. Fonov and Jussi Tohka for their guidance.

## Author contributions

Conceptualization: J.D.L., M.N.C. and B.T.K.; Data acquisition: A.A.M., M.S., H.M.B., A.D., K.R., L.G.L.; Data analysis: J.D.L.; Writing – original draft: J.D.L., M.N.C. and B.T.K.; Writing - review & editing: all authors.

