## Supplementary Material for "Neuroprognostication via Spatially-Informed Machine Learning Following Hypoxic-Ischemic Injury"

- eFigure 1: The population-specific neonatal brain multi-contrast template.
- eTable 1: The demographic, clinical, and laboratory data for the neonates used in the Bayley cognitive outcomes prediction analyses.
- eTable 2: The demographic, clinical, and laboratory data for the neonates used in the Bayley expressive language outcomes prediction analyses.
- eTable 3: The demographic, clinical, and laboratory data for the neonates used in the Bayley receptive language outcomes prediction analyses.
- eTable 4: The demographic, clinical, and laboratory data for the neonates used in the Bayley gross motor outcomes prediction analyses.
- eTable 5: The demographic, clinical, and laboratory data for the neonates used in the Bayley fine motor outcomes prediction analyses.

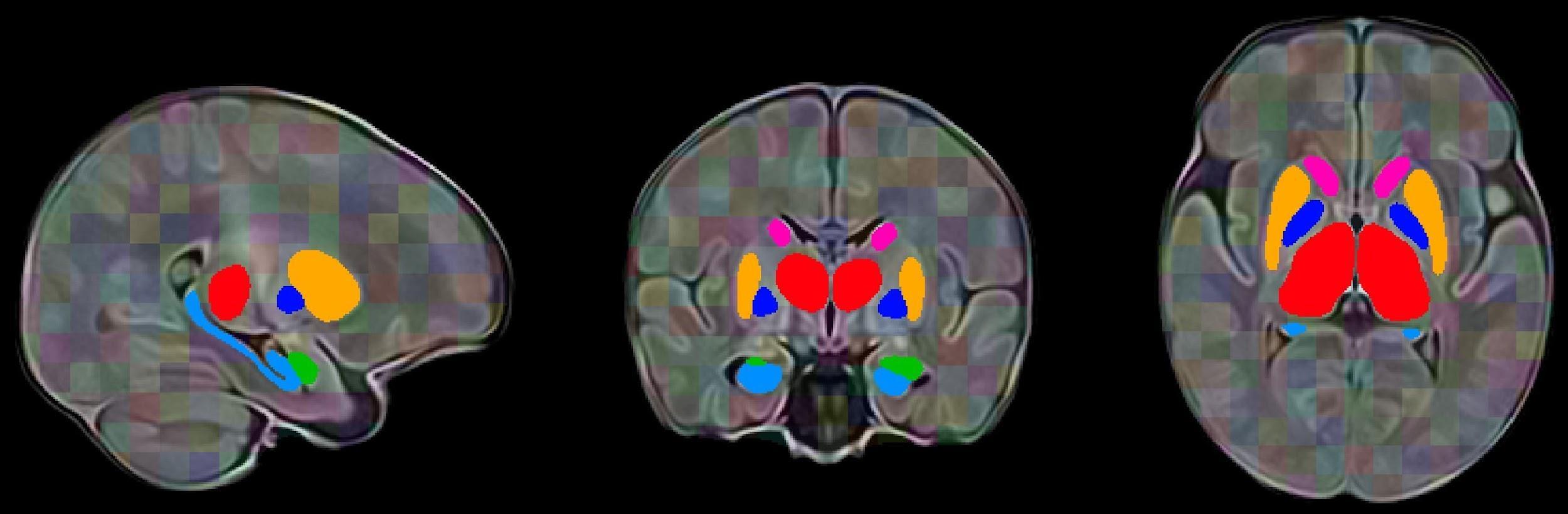

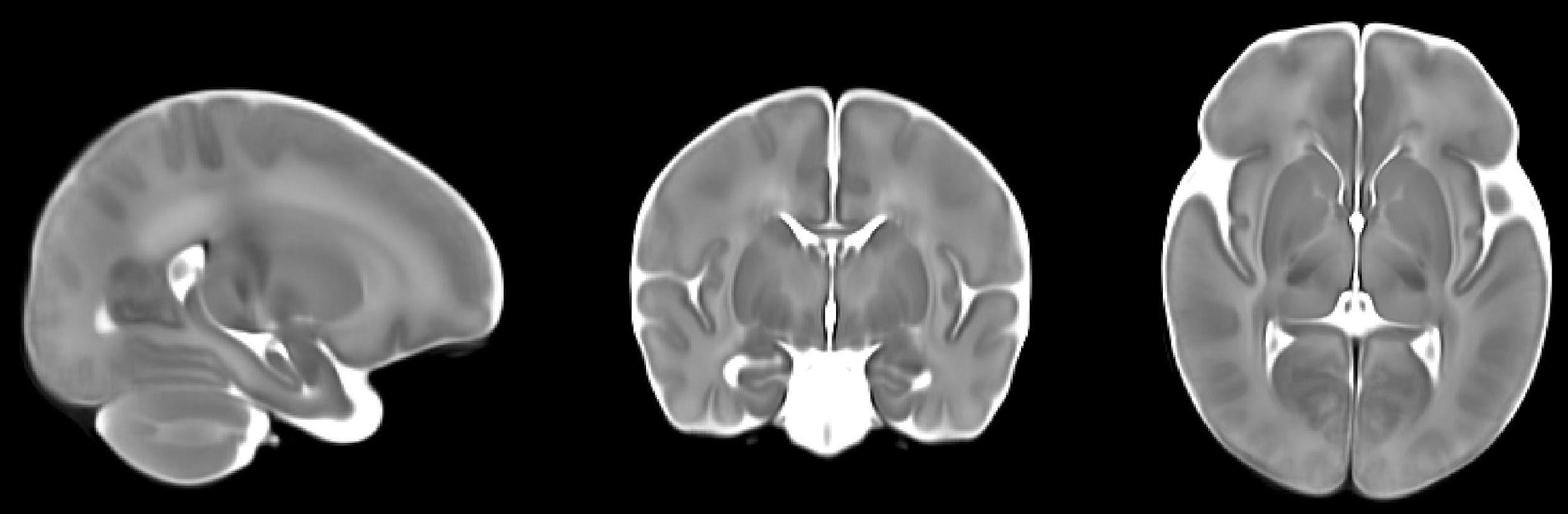
**e**
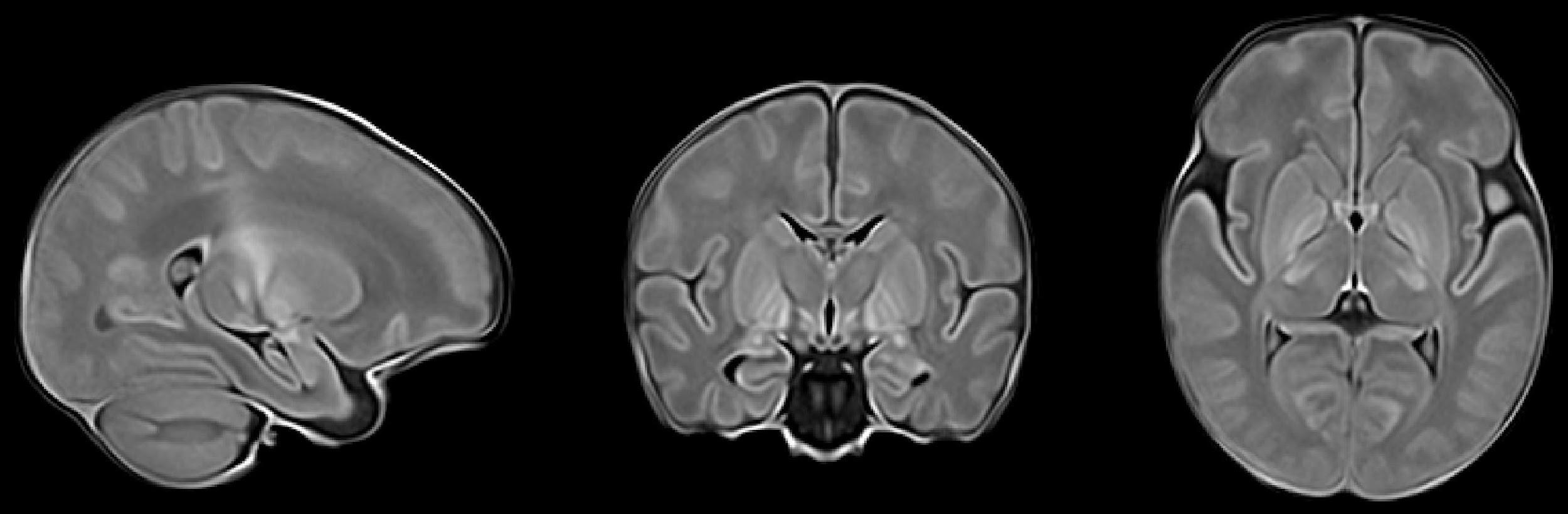
**Figure 1**: **The population-specific neonatal brain multi-contrast template.** The top row shows the T1-weighted volume ; the second row shows the T2-weighted volume ; the bottom row shows the T1-weighted volume with the labels for the amygdala, hippocampus, the subcortical gray-matter structures, and the cubic tiling of the entire brain overlaid on it. The amygdala is shown in green ; the hippocampus in light blue ; the globus pallidus in dark blue ; the putamen in gold ; the caudate in pink ; the thalamus in red ; and the cubic tiles in random translucent colors.

**eTable 1: The demographic, clinical, and laboratory data for the neonates used in the Bayley cognitive outcomes prediction analyses.** The median (x˜), mean (*µ*), and standard deviation (*σ*) are presented for males and females, separately. The number of values available are presented in parentheses.

|  | **Female** | **Male** |
| --- | --- | --- |
|  | x˜ , *µ ± σ* (*N* ) | x˜ , *µ ± σ* (*N* ) |
| **Bayley cognitive, percentile** | 50.00 , 44*.*31 *±* 27*.*21 (68) | 37.00 , 42*.*05 *±* 25*.*82 (100) |
| **Gestational age, weeks** | 39.29 , 39*.*11 *±* 1*.*47 (68) | 39.14 , 38*.*90 *±* 1*.*63 (100) |
| **Birth weight, g** | 3182.50 , 3186*.*50 *±* 548*.*04 (68) | 3340.00 , 3357*.*95 *±* 573*.*86 (100) |
| **5-minute Apgar score** | 5.00 , 4*.*58 *±* 2*.*24 (62) | 5.00 , 4*.*69 *±* 1*.*95 (96) |
| **Cord venous pH** | 7.16 , 7*.*13 *±* 0*.*15 (60) | 7.13 , 7*.*10 *±* 0*.*15 (90) |
| **Cord arterial pH** | 7.03 , 7*.*03 *±* 0*.*15 (60) | 6.99 , 7*.*01 *±* 0*.*15 (92) |
| **First postnatal pH** | 7.14 , 7*.*12 *±* 0*.*15 (63) | 7.10 , 7*.*10 *±* 0*.*15 (87) |
| **Highest lactate, mmol/l** | 8.20 , 8*.*82 *±* 4*.*70 (67) | 8.15 , 8*.*99 *±* 5*.*08 (100) |
| **Highest Thompson score** | 8.50 , 8*.*74 *±* 4*.*11 (58) | 8.00 , 8*.*62 *±* 3*.*38 (87) |

**eTable 2: The demographic, clinical, and laboratory data for the neonates used in the Bayley expressive language outcomes prediction analyses.** The median (x˜), mean (*µ*), and standard deviation (*σ*) are presented for males and females, separately. The number of values available are presented in parentheses.

|  | **Female** | **Male** |
| --- | --- | --- |
|  | x˜ , *µ ± σ* (*N* ) | x˜ , *µ ± σ* (*N* ) |
| **Bayley expr. lang., percentile** | 31.00 , 37*.*24 *±* 29*.*96 (58) | 20.50 , 28*.*16 *±* 26*.*52 (90) |
| **Gestational age, weeks** | 39.57 , 39*.*17 *±* 1*.*54 (58) | 39.14 , 38*.*89 *±* 1*.*67 (90) |
| **Birth weight, g** | 3192.50 , 3192*.*12 *±* 557*.*61 (58) | 3330.00 , 3352*.*46 *±* 596*.*59 (90) |
| **5-minute Apgar score** | 5.00 , 4*.*47 *±* 2*.*30 (53) | 5.00 , 4*.*62 *±* 1*.*98 (86) |
| **Cord venous pH** | 7.15 , 7*.*13 *±* 0*.*15 (53) | 7.14 , 7*.*10 *±* 0*.*15 (80) |
| **Cord arterial pH** | 7.03 , 7*.*03 *±* 0*.*14 (52) | 6.99 , 7*.*01 *±* 0*.*15 (82) |
| **First postnatal pH** | 7.14 , 7*.*12 *±* 0*.*15 (53) | 7.10 , 7*.*09 *±* 0*.*15 (78) |
| **Highest lactate, mmol/l** | 8.20 , 8*.*90 *±* 4*.*70 (57) | 7.85 , 8*.*62 *±* 5*.*00 (90) |
| **Highest Thompson score** | 9.00 , 8*.*92 *±* 3*.*95 (48) | 8.00 , 8*.*46 *±* 3*.*24 (78) |

**eTable 3: The demographic, clinical, and laboratory data for the neonates used in the Bayley receptive language outcomes prediction analyses.** The median (x˜), mean (*µ*), and standard deviation (*σ*) are presented for males and females, separately. The number of values available are presented in parentheses.

|  | **Female** | **Male** |
| --- | --- | --- |
|  | x˜ , *µ ± σ* (*N* ) | x˜ , *µ ± σ* (*N* ) |
| **Bayley recept. lang., percentile** | 50.00 , 47*.*09 *±* 30*.*91 (58) | 37.00 , 36*.*33 *±* 27*.*58 (90) |
| **Gestational age, weeks** | 39.43 , 39*.*12 *±* 1*.*53 (58) | 39.14 , 38*.*89 *±* 1*.*67 (90) |
| **Birth weight, g** | 3182.50 , 3159*.*53 *±* 540*.*14 (58) | 3330.00 , 3352*.*46 *±* 596*.*59 (90) |
| **5-minute Apgar score** | 5.00 , 4*.*45 *±* 2*.*28 (53) | 5.00 , 4*.*62 *±* 1*.*98 (86) |
| **Cord venous pH** | 7.15 , 7*.*13 *±* 0*.*15 (53) | 7.14 , 7*.*10 *±* 0*.*15 (80) |
| **Cord arterial pH** | 7.02 , 7*.*03 *±* 0*.*14 (51) | 6.99 , 7*.*01 *±* 0*.*15 (82) |
| **First postnatal pH** | 7.16 , 7*.*12 *±* 0*.*15 (53) | 7.10 , 7*.*09 *±* 0*.*15 (78) |
| **Highest lactate, mmol/l** | 8.20 , 8*.*90 *±* 4*.*70 (57) | 7.85 , 8*.*62 *±* 5*.*00 (90) |
| **Highest Thompson score** | 8.50 , 8*.*77 *±* 4*.*04 (48) | 8.00 , 8*.*46 *±* 3*.*24 (78) |

**eTable 4: The demographic, clinical, and laboratory data for the neonates used in the Bayley gross motor outcomes prediction analyses.** The median (x˜), mean (*µ*), and standard deviation (*σ*) are presented for males and females, separately. The number of values available are presented in parentheses.

|  | **Female** | **Male** |
| --- | --- | --- |
|  | x˜ , *µ ± σ* (*N* ) | x˜ , *µ ± σ* (*N* ) |
| **Bayley gross mtr., percentile** | 37.00 , 40*.*44 *±* 19*.*06 (62) | 37.00 , 41*.*90 *±* 20*.*79 (93) |
| **Gestational age, weeks** | 39.36 , 39*.*13 *±* 1*.*49 (62) | 39.14 , 38*.*90 *±* 1*.*67 (93) |
| **Birth weight, g** | 3192.50 , 3206*.*73 *±* 547*.*20 (62) | 3360.00 , 3377*.*58 *±* 577*.*17 (93) |
| **5-minute Apgar score** | 5.00 , 4*.*52 *±* 2*.*26 (56) | 5.00 , 4*.*64 *±* 1*.*96 (89) |
| **Cord venous pH** | 7.15 , 7*.*12 *±* 0*.*15 (54) | 7.14 , 7*.*10 *±* 0*.*15 (83) |
| **Cord arterial pH** | 7.00 , 7*.*02 *±* 0*.*15 (55) | 6.99 , 7*.*01 *±* 0*.*15 (85) |
| **First postnatal pH** | 7.14 , 7*.*11 *±* 0*.*16 (57) | 7.09 , 7*.*09 *±* 0*.*15 (81) |
| **Highest lactate, mmol/l** | 8.30 , 9*.*03 *±* 4*.*78 (61) | 7.90 , 8*.*74 *±* 5*.*05 (93) |
| **Highest Thompson score** | 9.00 , 9*.*11 *±* 4*.*06 (53) | 8.00 , 8*.*57 *±* 3*.*29 (81) |

**eTable 5: The demographic, clinical, and laboratory data for the neonates used in the Bayley fine motor outcomes prediction analyses.** The median (x˜), mean (*µ*), and standard deviation (*σ*) are presented for males and females, separately. The number of values available are presented in parentheses.

|  | **Female** | **Male** |
| --- | --- | --- |
|  | x˜ , *µ ± σ* (*N* ) | x˜ , *µ ± σ* (*N* ) |
| **Bayley fine motor, percentile** | 63.00 , 53*.*83 *±* 26*.*06 (65) | 50.00 , 49*.*91 *±* 26*.*42 (94) |
| **Gestational age, weeks** | 39.29 , 39*.*11 *±* 1*.*48 (65) | 39.14 , 38*.*88 *±* 1*.*67 (94) |
| **Birth weight, g** | 3185.00 , 3195*.*49 *±* 555*.*54 (65) | 3350.00 , 3367*.*71 *±* 581*.*98 (94) |
| **5-minute Apgar score** | 5.00 , 4*.*58 *±* 2*.*21 (59) | 5.00 , 4*.*64 *±* 1*.*94 (90) |
| **Cord venous pH** | 7.15 , 7*.*13 *±* 0*.*15 (57) | 7.14 , 7*.*10 *±* 0*.*15 (84) |
| **Cord arterial pH** | 7.00 , 7*.*02 *±* 0*.*15 (57) | 6.99 , 7*.*01 *±* 0*.*15 (86) |
| **First postnatal pH** | 7.16 , 7*.*12 *±* 0*.*16 (60) | 7.10 , 7*.*09 *±* 0*.*15 (82) |
| **Highest lactate, mmol/l** | 8.20 , 8*.*84 *±* 4*.*74 (64) | 8.00 , 8*.*79 *±* 5*.*04 (94) |
| **Highest Thompson score** | 9.00 , 8*.*98 *±* 4*.*07 (55) | 8.00 , 8*.*59 *±* 3*.*27 (82) |
